# Efficacy of Testosterone Replacement Therapy in Men With Hypogonadism: A Systematic Review and Meta-Analysis of Clinical Outcomes, Body Composition, and Quality of Life

**DOI:** 10.64898/2026.09.27.26364129

**Authors:** Thiago de Sousa Siqueira, Hércules Kanaan Pereira Sousa, Ricardo Rezende de Queiroz

**Affiliations:** Centro Universitário UNINOVAFAPI

**Keywords:** Hypogonadism, Testosterone, Testosterone replacement therapy, Meta-analysis, Men’s health

## Abstract

**Background:** Male hypogonadism is associated with sexual, physical, and psychological manifestations, and testosterone replacement therapy (TRT) is used to restore testosterone levels and relieve symptoms.

**Objective:** To evaluate the efficacy of TRT in adult men with hypogonadism across erectile function, hypogonadal symptoms, fatigue, body composition, waist circumference, urinary symptoms, and quality of life.

**Methods:** This systematic review and meta-analysis followed PRISMA guidance. PubMed/MEDLINE, Google Scholar, and ScienceDirect were searched. Fourteen studies involving 1,388 participants were included. Effect estimates were synthesized as standardized mean differences using random-effects models, and heterogeneity was assessed with I^2^, Cochran’s Q, and τ^2^.

**Results:** TRT was associated with favorable pooled effects on erectile function, hypogonadal symptoms, fatigue, lean mass, fat mass, waist circumference, and quality of life. No consistent effect was observed for lower urinary tract symptoms. Heterogeneity was high for erectile function, hypogonadal symptoms, waist circumference, and urinary symptoms; moderate for body-composition outcomes; and low for fatigue and quality of life. Randomized trials generally yielded smaller effect estimates than observational studies.

**Conclusions:** TRT was associated with improvement across several clinical and body-composition outcomes in hypogonadal men, although effect magnitude varied substantially by outcome and study design. The high heterogeneity for several outcomes and the larger estimates in observational studies warrant cautious interpretation and support the need for longer, high-quality randomized trials.

## Introduction

Male hypogonadism is a clinical and biochemical syndrome characterized by inadequate testosterone production and may be classified as primary (testicular failure) or secondary (hypothalamic-pituitary dysfunction). Its prevalence increases with age, affecting approximately 20% of men older than 60 years and up to 50% of men with comorbidities such as diabetes and obesity. Despite its frequency, hypogonadism remains underdiagnosed in clinical practice because its symptoms overlap with normal aging and screening uptake is limited. The impact on quality of life can be substantial, affecting physical, sexual, and psychological functioning (Silva et al., 2024; Giannetta et al., 2012; Grant; Anawalt, 2003; Marcelli; Mediwala, 2020; Seftel, 2006).

The clinical manifestations of hypogonadism span multiple domains. Sexual manifestations include erectile dysfunction, reduced libido, and diminished orgasmic pleasure. Physical manifestations include chronic fatigue, reduced muscle mass and strength, increased visceral adiposity, and osteopenia. Psychological manifestations include irritability, depression, anxiety, and reduced well-being. Together, these symptoms may adversely affect work capacity, interpersonal relationships, and functional independence, particularly in older adults and in men with associated comorbidities (Dandona; Rosenberg, 2010; Indirli et al., 2023; Kumar et al., 2010; Marcelli; Mediwala, 2020).

Hypogonadism is associated with clinically relevant comorbidities that may worsen prognosis. Testosterone deficiency and metabolic syndrome have a bidirectional relationship, and hypogonadism is common among men with type 2 diabetes. Central obesity, particularly visceral adiposity, is inversely correlated with serum testosterone concentrations, creating a cycle of worsening metabolic dysfunction. Epidemiological studies have also associated low testosterone levels with cardiovascular events. In addition, androgen deficiency adversely affects bone health, increasing the risk of osteoporosis and fragility fractures, especially in older men (Dandona; Rosenberg, 2010; Dimopoulou et al., 2018; Yeo et al., 2021; Zarotsky et al., 2014).

Testosterone exerts biological effects through androgen receptors expressed in multiple target tissues, including skeletal muscle, adipose tissue, the vascular endothelium, and the central nervous system. In skeletal muscle, androgen-receptor activation promotes gene transcription and protein synthesis, thereby supporting hypertrophy and strength. In adipose tissue, testosterone activates lipolytic pathways and may reduce visceral fat accumulation. Testosterone also modulates endothelial function through nitric oxide bioavailability and influences dopaminergic and serotonergic neurotransmission in brain regions involved in libido, mood, and energy. These mechanisms provide the biological rationale for testosterone replacement in men with hypogonadism (Lopes et al., 2012; Mudali; Dobs, 2004; Sheffield-Moore, 2000; Zitzmann, 2009).

This review was motivated by the need for a broad meta-analysis evaluating multiple clinical outcomes of testosterone replacement therapy (TRT) simultaneously, including erectile function, hypogonadal symptoms, fatigue, body composition, waist circumference, urinary symptoms, and quality of life, while considering differences between randomized clinical trials and observational studies. The primary objective was to evaluate the efficacy of TRT in hypogonadal men for each outcome. Secondary objectives were to compare effect estimates across study designs, quantify between-study heterogeneity, and explore potential sources of heterogeneity to provide an integrated summary of the available evidence.

## Methods

### Study design and registration

This systematic review was conducted in accordance with the Preferred Reporting Items for Systematic Reviews and Meta-Analyses (PRISMA) guidance. The protocol was prospectively registered in the Open Science Framework (OSF). The search, study selection, data extraction, and analyses were conducted between January and April 2026 according to the prespecified protocol.

### Eligibility criteria

Studies were eligible if they included adult men (>18 years) with confirmed hypogonadism, defined as total testosterone <12 nmol/L or an equivalent criterion based on free testosterone, who received testosterone replacement by any route (intramuscular, transdermal, sublingual, or oral). Eligible comparators were placebo, standard treatment, an untreated group, or no comparator. Outcomes of interest were erectile function, hypogonadal symptoms, fatigue, lean mass, fat mass, waist circumference, urinary symptoms, and quality of life. Randomized clinical trials, nonrandomized controlled trials, and observational studies with or without a control group were eligible. Animal studies, case reports, case series, and reviews without primary data were excluded.

### Search strategy

PubMed/MEDLINE, Google Scholar, and ScienceDirect were searched for studies published from January 1990 through April 2026. The search strategy combined MeSH terms and free-text descriptors related to testosterone (testosterone replacement therapy, TRT, androgen therapy), hypogonadism (hypogonadism, late-onset hypogonadism, testosterone deficiency), and the outcomes of interest (erectile function, IIEF, muscle mass, body composition, fatigue, quality of life). Reference lists of included articles were also screened manually, and Google Scholar was used to search for gray literature, including unpublished studies and conference proceedings.

### Study selection

Two reviewers independently selected studies in two stages: title and abstract screening followed by full-text assessment of potentially eligible articles. Disagreements were resolved by consensus; when consensus could not be reached, a third reviewer made the final decision. The PRISMA flow diagram showing the numbers of records identified, screened, assessed for eligibility, and included is provided as Figure S1 in the Supplementary Material. Zotero was used for reference management and organization of the selection process.

### Data extraction

Two reviewers independently extracted data using a standardized, pretested form. Extracted variables included first author, publication year, country, study design, sample size, age, diagnostic criteria for hypogonadism, intervention characteristics (dose, route of administration, and treatment duration), control group, outcomes, and standardized mean difference (SMD) and standard error (SE) when reported directly or when calculable from means, standard deviations, confidence intervals, or p values. Extraction disagreements were resolved by consensus, with third-reviewer arbitration when necessary.

### Risk of bias assessment

Two reviewers independently assessed risk of bias. Randomized clinical trials were evaluated using the Cochrane Risk of Bias 2 (RoB 2) tool across the domains of randomization, deviations from intended interventions, missing outcome data, outcome measurement, and selective reporting. Nonrandomized studies were assessed using ROBINS-I. Risk of bias was classified as low, moderate, high, or critical according to the algorithms of the respective tools. Disagreements were resolved by consensus. Studies judged to have critical risk of bias were excluded from the primary analyses.

### Statistical analysis

The primary effect measure was the standardized mean difference (SMD). Random-effects models were fitted using the restricted maximum likelihood (REML) estimator because moderate to high clinical and methodological heterogeneity was expected. The SE of the SMD was extracted or calculated from the source articles. When SMD and SE were not directly available, they were derived from baseline and final means and standard deviations for pre-post studies, between-group differences in change, 95% confidence intervals, or p values, with Hedges’ correction for small samples. The Knapp-Hartung adjustment was applied to mixed-effects models to adjust degrees of freedom and obtain more conservative confidence intervals. For dichotomous outcomes expressed as proportions, an arcsine square-root transformation was used before conversion to the SMD metric. Analyses were performed in JASP, with selected results validated in R using the metafor package. Results are reported as SMDs with 95% confidence intervals (95% CIs).

Between-study heterogeneity was evaluated using I^2^ (classified as low, <40%; moderate, 40%-75%; and high, >75%), Cochran’s Q with its associated p value, and τ^2^ as the between-study variance. High I^2^ values were interpreted as substantial heterogeneity and supported the use of random-effects models and sensitivity analyses.

## Results

### Study selection and characteristics

The study identification, screening, eligibility assessment, and inclusion process is summarized in the PRISMA flow diagram (Figure S1, Supplementary Material). A total of 11,094 records were identified, including 11,050 records from databases and 44 from registers. After removal of 1,250 duplicate records, 9,844 records underwent title and abstract screening, of which 9,600 were excluded. Subsequently, 244 reports were sought for retrieval; four could not be retrieved, leaving 240 full-text reports assessed for eligibility. Of these, 226 were excluded: 78 did not meet the diagnostic criteria for hypogonadism or had insufficient biochemical data, 62 did not report the outcomes of interest or had incomplete data, 51 had an ineligible study design, and 35 had a population, intervention, or comparator outside the eligibility criteria. Fourteen studies were ultimately included in the systematic review and meta-analysis (Figure S1).

TRT was administered by several routes, including intramuscular testosterone (enanthate or undecanoate, including regimens of 1,000 mg every 6-12 weeks), transdermal testosterone (1%-2% gel, 25-100 mg/day), and sublingual formulations. Treatment duration ranged from 16 weeks to 11 years, with a median follow-up of approximately 12 months in most studies.

Comparators varied across studies and included placebo, untreated groups, and comparisons between different testosterone formulations. Some observational studies used pre-post analyses without a control group. This methodological heterogeneity was considered when interpreting the pooled estimates.

Across the included studies, 1,388 participants were randomized or allocated to intervention groups, with individual study sample sizes ranging from 20 to 262 participants. Overall, risk of bias was lower in double-blind placebo-controlled studies, intermediate in studies with nonblinded control groups, and higher in uncontrolled observational studies, mainly because of the absence of randomization and the potential influence of uncontrolled placebo effects, regression to the mean, and confounding.

### Erectile function

TRT was associated with improvement in erectile function, assessed using erectile-function domains of the International Index of Erectile Function or the total IIEF-5 score. The pooled effect from 12 studies favored TRT (SMD = 1.23; 95% CI, 0.50 to 1.96; p = 0.003), although heterogeneity was very high (I^2^ = 97.8%), reflecting differences in study design and populations (Figure S4).

When considered separately, double-blind placebo-controlled randomized trials generally showed modest but consistent effects of testosterone on erectile function. In these methodologically stronger studies, effect sizes ranged from small to moderate. Larger effects were observed in populations with more marked hypogonadism and in participants with comorbidities such as type 2 diabetes. By contrast, uncontrolled observational and pre-post studies produced substantially larger effect estimates, suggesting that design-related biases, including regression to the mean and uncontrolled nonspecific effects, may contribute to the larger observed benefits.

The very high heterogeneity indicates that the estimated effects were not uniform across populations and study designs. This variability limits the precision and generalizability of the pooled estimate and warrants cautious interpretation.

From a clinical perspective, the improvement in erectile function was more modest in the higher-quality randomized evidence than in the observational evidence. Benefits appeared more evident in men with unequivocal hypogonadism and pre-existing erectile dysfunction. The variability across studies underscores the importance of individualized clinical assessment and realistic expectations regarding the magnitude of improvement.

### Hypogonadal symptoms

TRT improved hypogonadal symptoms measured with the Aging Males’ Symptoms (AMS) scale. The pooled effect from nine studies favored TRT (SMD = 1.56; 95% CI, 0.59 to 2.53; p = 0.006), with extremely high heterogeneity (I^2^ = 98.6%) (Figure S8).

Across AMS domains, the largest improvements were reported in somatic and sexual domains, including physical energy, sleep quality, vitality, libido, and sexual performance. Psychological symptoms such as irritability, anxiety, and depressed mood showed smaller and less consistent responses across studies.

As with erectile function, placebo-controlled randomized trials generally yielded small-to-moderate effects on hypogonadal symptoms, whereas uncontrolled observational and pre-post studies produced substantially larger estimates, in some cases more than twice those seen in randomized trials. Placebo effects, regression to the mean, and measurement bias may therefore have contributed to larger estimates in studies with lower methodological control.

Potential sources of heterogeneity included baseline severity of hypogonadism, comorbid diabetes and obesity, treatment duration ranging from 16 weeks to more than five years, route of testosterone administration, and differences in symptom-assessment instruments. Baseline depression attenuated the response to TRT in some studies and may represent an additional source of between-study variability.

### Fatigue and energy

TRT was associated with reduced fatigue and increased energy. The pooled analysis of three studies favored TRT (SMD = 0.42; 95% CI, 0.15 to 0.68; p = 0.021), with low heterogeneity (I^2^ = 0.0%) (Figure S3).

Unlike several other outcomes, the studies assessing fatigue showed relatively homogeneous results despite differences in populations and assessment instruments. The direction of effect was consistent, with reduced tiredness and general fatigue accompanied by improvements in energy and vitality. Benefits were observed in both shorter and longer studies, suggesting that improvements in fatigue may emerge relatively early and persist during continued treatment.

### Lean mass and muscle strength

TRT increased lean mass and was associated with improvements in muscle strength. The pooled analysis of six studies favored TRT (SMD = 0.62; 95% CI, 0.27 to 0.97; p = 0.006), with moderate-to-high heterogeneity (I^2^ = 73.6%) (Figure S6).

The magnitude of lean-mass gain varied by dose, route, and treatment duration and tended to be greater in studies using higher-dose parenteral testosterone and in men with more severe hypogonadism. Increases in skeletal muscle mass, particularly in the lower limbs, were accompanied in some studies by functional improvements such as greater leg-press strength.

These changes may be clinically relevant in vulnerable populations, including frail older adults and patients with chronic debilitating disease. In men with obesity undergoing weight-loss interventions, TRT was also reported to attenuate loss of lean mass during caloric restriction.

### Fat mass

TRT was associated with reduced fat mass and body-fat percentage. The pooled analysis of four studies favored TRT (SMD = 0.57; 95% CI, 0.17 to 0.98; p = 0.021), with moderate heterogeneity (I^2^ = 58.7%) (Figure S5).

The magnitude of fat-mass reduction varied according to testosterone dose and treatment duration and was more pronounced in studies using higher doses and longer follow-up. Reported fat loss occurred predominantly in the trunk compartment, which includes metabolically relevant visceral adiposity.

Reductions in adiposity were accompanied in some studies by improvements in metabolic parameters, including triglycerides and waist circumference. These findings suggest that body-composition changes may contribute to a more favorable cardiometabolic profile in hypogonadal men, particularly those with obesity or metabolic syndrome.

### Waist circumference

TRT reduced waist circumference, a marker of central adiposity and a component of metabolic syndrome. The pooled analysis of four studies favored TRT (SMD = 0.69; 95% CI, 0.31 to 1.07; p = 0.011), with high heterogeneity (I^2^ = 76.1%) (Figure S2).

Reductions in waist circumference were reported in both shorter and longer studies. Effects tended to be more pronounced in populations with greater baseline obesity and in men with established metabolic syndrome. Because waist circumference is associated with cardiometabolic risk, this finding may be clinically relevant, although the high heterogeneity limits the precision of the pooled estimate.

### Lower urinary tract symptoms

TRT did not show a consistent effect on lower urinary tract symptoms assessed with the International Prostate Symptom Score (IPSS). The pooled analysis of four studies was not statistically significant (SMD = 0.61; 95% CI, -0.43 to 1.65; p = 0.159), and heterogeneity was extremely high (I^2^ = 96.5%) (Figure S9).

Results differed markedly across studies. One propensity score-matched observational study reported a large favorable effect on urinary symptoms, whereas the remaining studies showed small-to-moderate, nonsignificant, or null effects.

Potential explanations for this heterogeneity include differences in baseline urinary-symptom severity, the prevalence of benign prostatic hyperplasia, and study design. The predominance of observational evidence for some estimates may also have contributed to larger apparent benefits. The prostate saturation model provides one possible biological explanation for the absence of a consistent worsening of urinary symptoms once testosterone concentrations are restored to the physiological range.

### Quality of life

TRT was associated with improved overall quality of life in hypogonadal men. The pooled analysis of three studies favored TRT (SMD = 0.56; 95% CI, 0.17 to 0.94; p = 0.025), with low heterogeneity (I^2^ = 0.0%) (Figure S7).

The instruments used to assess quality of life captured different dimensions. The physical component of the Short Form-36 improved in some studies, reflecting gains in functional capacity, energy for daily activities, and perceived general health. Global treatment-response measures and subjective well-being also favored testosterone in the included evidence.

Quality of life is an integrative patient-centered outcome that may reflect changes across several domains, including physical function, energy, mood, sexual function, and body composition. The low heterogeneity observed for this outcome provides greater consistency than was seen for several other endpoints.

### Summary of pooled effects

Table 1 summarizes the direction of the pooled TRT effect and heterogeneity for each outcome. Heterogeneity was high for erectile function, hypogonadal symptoms, waist circumference, and urinary symptoms; moderate for lean mass and fat mass; and low for fatigue and quality of life.

**Table 1.** Summary of TRT effects by outcome.

| Outcome | Direction of pooled effect | Heterogeneity (I<sup>2</sup>) |
| --- | --- | --- |
| Erectile function | Favorable | High (97.8%) |
| Hypogonadal symptoms (AMS) | Favorable | High (98.6%) |
| Fatigue and energy | Favorable | Low (0.0%) |
| Lean mass | Favorable | Moderate (73.6%) |
| Fat mass | Favorable | Moderate (58.7%) |
| Waist circumference | Favorable | High (76.1%) |
| Urinary symptoms (IPSS) | Inconclusive | High (96.5%) |
| Quality of life | Favorable | Low (0.0%) |

Source: Authors, 2026.

## Discussion

### Principal findings

This meta-analysis found that TRT was associated with improvement across multiple clinical outcomes in men with hypogonadism. Body-composition outcomes showed favorable pooled effects, including increased lean mass and reduced fat mass and waist circumference, with concurrent improvements in muscle strength reported in some studies. Fatigue and energy improved with low between-study heterogeneity, and quality of life also showed a favorable pooled effect. Erectile function and hypogonadal symptoms improved overall but showed very high heterogeneity. Lower urinary tract symptoms were the only outcome without a consistent pooled effect. Across outcomes, randomized trials generally produced smaller estimates than observational studies, highlighting the influence of study design on apparent treatment magnitude.

### Comparison with previous evidence and study design

The findings are broadly consistent with previous systematic reviews and meta-analyses of TRT in hypogonadal men. Earlier work has reported favorable effects on lean mass, fat mass, bone-related outcomes, erectile function, and hypogonadal symptoms, although effect size and certainty have varied across reviews (Elliott et al., 2017; Guo et al., 2016; Madsen et al., 2022; Ponce et al., 2018).

Two higher-quality randomized trials in the included evidence did not show significant benefit for specific outcomes. Paduch and colleagues, who evaluated ejaculatory dysfunction, found no significant testosterone effect compared with placebo. Højer and colleagues, studying testicular-cancer survivors with mild Leydig-cell insufficiency, likewise reported no improvement in sexual function or quality of life. These studies suggest that men with milder biochemical deficiency or specific etiologies may respond differently to testosterone replacement.

A notable finding was the discrepancy between double-blind placebo-controlled randomized trials and observational or pre-post studies. Observational studies consistently yielded substantially larger effect estimates, especially for erectile function and hypogonadal symptoms. In methodologically stronger randomized trials, effect sizes were generally small to moderate. This contrast emphasizes the importance of considering study design when interpreting pooled estimates.

Several biases may contribute to larger estimates in uncontrolled studies. Placebo and expectancy effects may be particularly relevant for subjective outcomes such as sexual function, mood, and quality of life. Regression to the mean can also inflate apparent improvement in pre-post designs, and publication bias may favor the publication of positive findings (Boloña et al., 2007; Corona et al., 2017; Elliott et al., 2017; Isidori et al., 2005; Walther; Breidenstein; Miller, 2019; Yang et al., 2023). These issues support reporting randomized and observational evidence separately when both designs are included in evidence syntheses.

### Biological mechanisms

The observed effects are biologically plausible. In skeletal muscle, testosterone activates the nuclear androgen receptor, promotes transcription and protein synthesis, and supports hypertrophy, particularly of type II muscle fibers. Testosterone may also modulate myostatin, a negative regulator of muscle growth (Dandona et al., 2021; Ghanim et al., 2019; Kvorning et al., 2007; Oura et al., 2025).

In adipose tissue, testosterone may promote lipolysis, increase fatty-acid oxidation, and reduce lipid uptake. Improvements in insulin sensitivity and glucose tolerance reported in some studies may reflect both reduced visceral adiposity and direct effects on glucose transport pathways, including GLUT-4 signaling (Pergola, 2000; Kelly; Jones, 2013; Rao; Kelly; Jones, 2013).

Within the central nervous system, testosterone and its aromatized metabolite estradiol influence dopaminergic and serotonergic neurotransmission in regions involved in libido, motivation, energy, and mood. The reduction in fatigue observed in this meta-analysis may therefore reflect both central effects and peripheral changes in muscle energy metabolism (Mędraś; Brona; JóŹków, 2018; Tobiansky et al., 2018).

Aromatization of testosterone to estradiol is also important for skeletal and metabolic effects. Estradiol is a major regulator of male bone homeostasis and suppresses osteoclastic resorption; studies of TRT and bone outcomes have reported concomitant increases in estradiol in settings where bone density was preserved or improved (Corona et al., 2022; Finkelstein et al., 2016; Golds; Houdek; Arnason, 2017; Tenuta et al., 2025).

The prostate saturation model proposes that androgen receptors in prostate tissue become saturated at relatively low physiological testosterone concentrations. Once physiological concentrations are restored, further increases may have limited additional effects on prostate growth or urinary symptoms, providing a possible explanation for the absence of a consistent pooled effect on IPSS (Haider et al., 2018; Khera et al., 2011; Xu et al., 2024).

### Clinical implications

The pooled evidence suggests that TRT can improve erectile and sexual function in some men with hypogonadism, although the effect was more modest in higher-quality randomized trials. Benefits appeared more pronounced in men with unequivocal hypogonadism and pre-existing erectile dysfunction. Testosterone may influence sexual function centrally through libido and peripherally through nitric-oxide signaling in the corpus cavernosum. The cited literature also supports evaluating treatment response over several months and considering combination therapy with phosphodiesterase-5 inhibitors in selected men who remain symptomatic (Aversa et al., 2019; Corona; Maggi, 2022; Rizk et al., 2017).

TRT was also associated with improvements in hypogonadal symptoms, fatigue, and quality of life, particularly in somatic and sexual domains of the AMS scale. The presence of baseline depression may attenuate the response, emphasizing the need to evaluate and manage relevant comorbidities when assessing persistent symptoms.

The body-composition findings were among the more consistent clinical signals, with increased lean mass and reductions in fat mass and waist circumference. These changes may be relevant for hypogonadal men with obesity, metabolic syndrome, or type 2 diabetes. However, TRT should be considered within established clinical indications and alongside lifestyle and risk-factor management rather than as a stand-alone treatment for obesity or metabolic disease.

The pooled evidence did not support using TRT specifically to improve lower urinary tract symptoms. At the same time, the included studies did not show a consistent worsening of IPSS after testosterone replacement. Given the very high heterogeneity and the limited number of studies, urinary symptoms and prostate-related factors should continue to be monitored according to standard clinical practice rather than inferred from the pooled estimate alone.

### Heterogeneity and potential effect modifiers

Statistical heterogeneity was high for several outcomes, particularly erectile function, hypogonadal symptoms, waist circumference, and urinary symptoms. Likely clinical sources included differences in the etiology and severity of hypogonadism, age, diabetes and obesity, concomitant medications such as phosphodiesterase-5 inhibitors, testosterone dose and route, and treatment duration, which ranged from 16 weeks to 11 years.

Methodological differences were also important. Double-blind randomized trials generally produced smaller effect estimates than uncontrolled observational and pre-post studies. The presence or absence of placebo control, allocation concealment, and overall risk of bias may therefore have contributed substantially to between-study variation.

Random-effects models were used because a common underlying effect was not assumed. Nevertheless, very high I^2^ values indicate that pooled estimates summarize substantially different effects across studies and should not be interpreted as a single effect expected in every patient population.

Sensitivity analyses and subgroup comparisons by study design were used to explore heterogeneity. These analyses indicated that randomized trials yielded more conservative estimates and were less susceptible to confounding than observational studies. Future meta-analyses should continue to distinguish estimates by study design when heterogeneous evidence bases are combined.

Potential modifiers of treatment response included baseline depression, obesity, severity of testosterone deficiency, comorbid diabetes or metabolic syndrome, treatment duration, and achieved serum testosterone concentrations. Men with higher body mass index and lower baseline testosterone tended to show larger changes in lean mass and waist circumference, whereas men with mild or borderline hypogonadism showed smaller and less consistent responses. Age was not a consistent effect modifier in the included evidence.

Treatment duration may also influence response. Changes in lean mass and strength may require several months, whereas changes in sexual function and mood may emerge earlier. The included evidence did not establish additional clinical benefit from maintaining testosterone concentrations above the physiological range.

### Strengths and limitations

This review has several limitations. First, substantial heterogeneity reduced the precision of pooled estimates and limits their generalizability. Second, the methodological quality of the included studies was heterogeneous. Only a subset were double-blind placebo-controlled randomized trials; the remainder included open-label and observational designs, sometimes without control groups, which are more susceptible to confounding, placebo effects, and regression to the mean. These limitations are particularly important for subjective outcomes such as sexual function and quality of life.

The relatively short duration of many randomized trials (16-24 weeks) may be insufficient for outcomes that evolve over longer periods, including muscle-mass gain and broader body-composition changes. Outcome instruments also varied across studies, particularly for erectile function (IIEF-5, IIEF-EF, MSHQ-EjD-SF) and hypogonadal symptoms (AMS and other symptom or mood scales), complicating standardization.

Formal assessment of publication bias was limited by the small number of studies available for individual outcomes. Selective publication of positive findings therefore cannot be excluded. The results also should not be generalized to women, adolescents, or men with contraindications to TRT because these populations were not represented in the included evidence.

Strengths include the broad search across multiple databases, manual reference screening, inclusion of multiple clinically relevant outcomes, and explicit consideration of heterogeneity and study design. Comparing randomized with observational evidence highlighted the consistently larger effect estimates in studies with less methodological control. The review also integrates outcomes beyond sexual function, including lean mass, fat mass, waist circumference, urinary symptoms, fatigue, and quality of life.

### Future research

Long-term randomized clinical trials with adequate placebo control are needed to assess the durability of benefits and longer-term safety, including cardiovascular events and mortality. Studies focused on clinically relevant subgroups, including men with baseline depression, severe obesity, and type 2 diabetes, may help clarify heterogeneity in response.

Greater standardization of outcome instruments, including consistent use of validated erectile-function and hypogonadal-symptom measures, would improve future evidence synthesis. Research on predictors of response, including baseline testosterone, sex hormone-binding globulin, and androgen-receptor polymorphisms, may also support more individualized treatment. Pragmatic prospective studies and registries may complement randomized-trial evidence by evaluating effectiveness in routine clinical settings.

## Conclusions

TRT was associated with improved body composition, reduced fatigue, and favorable pooled effects on erectile function, hypogonadal symptoms, and quality of life in men with hypogonadism. The magnitude of benefit was generally smaller in randomized trials than in observational studies, and several outcomes showed substantial heterogeneity. These findings support cautious, individualized interpretation of expected benefits and highlight the need for longer, methodologically rigorous randomized trials.

## Data Availability

All data produced are available online at medRxiv

## Declarations

### Ethics statement

This systematic review and meta-analysis used aggregate data from previously published studies and did not involve the collection of new individual participant data. Therefore, institutional ethics committee approval and participant consent were not required.

### Funding

The authors received no specific funding for this work.

### Competing interests

The authors declare no financial or personal competing interests that could have inappropriately influenced the work reported in this manuscript.

### Data availability

The data analyzed in this review are contained within the manuscript and its Supplementary Material.

### Generative AI disclosure

ChatGPT (OpenAI) was used to assist with English-language translation and editorial formatting of the manuscript. All AI-assisted text was reviewed by the authors, who remain responsible for the accuracy, integrity, and originality of the work.

### Author contributions

Thiago de Sousa Siqueira: Conceptualization, Methodology, Supervision, Writing - Original Draft, Project Administration.

Hércules Kanaan Pereira Sousa: Formal Analysis, Investigation, Software, Visualization. Ricardo Rezende de Queiroz: Resources, Funding Acquisition, Writing - Review & Editing.

## Notes

### Competing Interest Statement

The authors have declared no competing interest.

