## SUPPLEMENTARY MATERIAL for "Efficacy of Testosterone Replacement Therapy in Men With Hypogonadism: A Systematic Review and Meta-Analysis of Clinical Outcomes, Body Composition, and Quality of Life"

**Figure S1.** PRISMA flow diagram of the identification, screening, eligibility assessment, and inclusion of studies in the systematic review and meta-analysis.

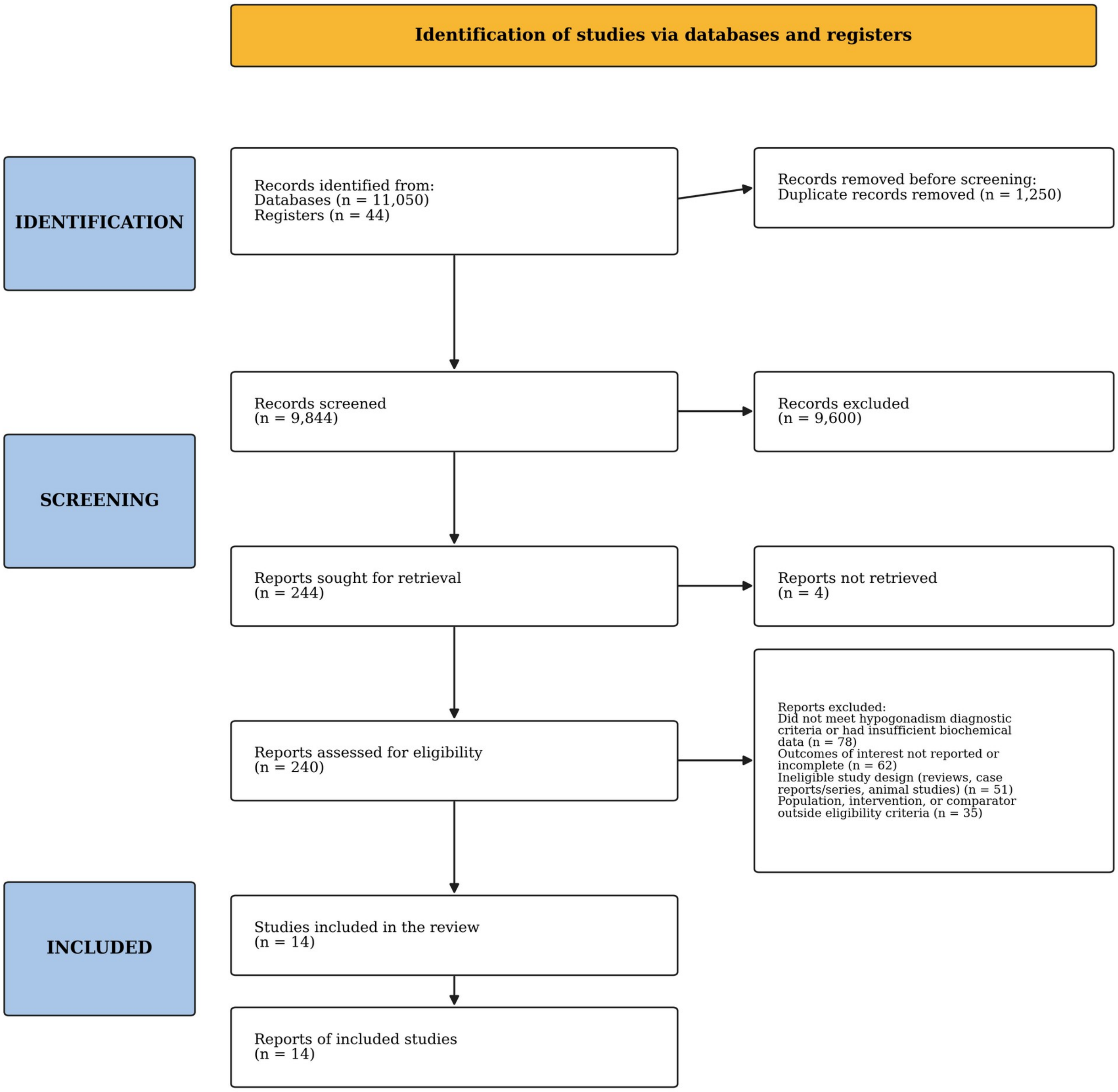


*Source: Authors, 2026.*

**Figure S2.** Forest plot of the effect of testosterone replacement therapy on waist circumference in men with hypogonadism.

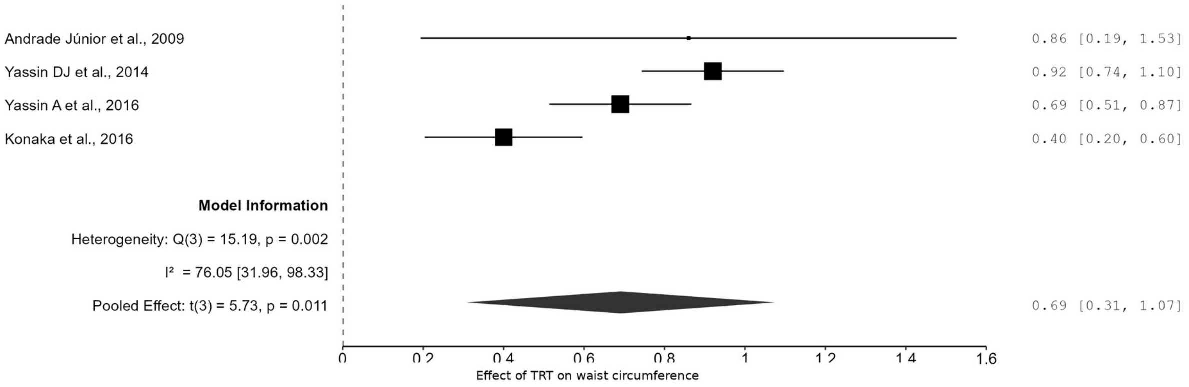


*Source: Authors, 2026.*

**Figure S3.** Forest plot of the effect of testosterone replacement therapy on fatigue in men with hypogonadism.

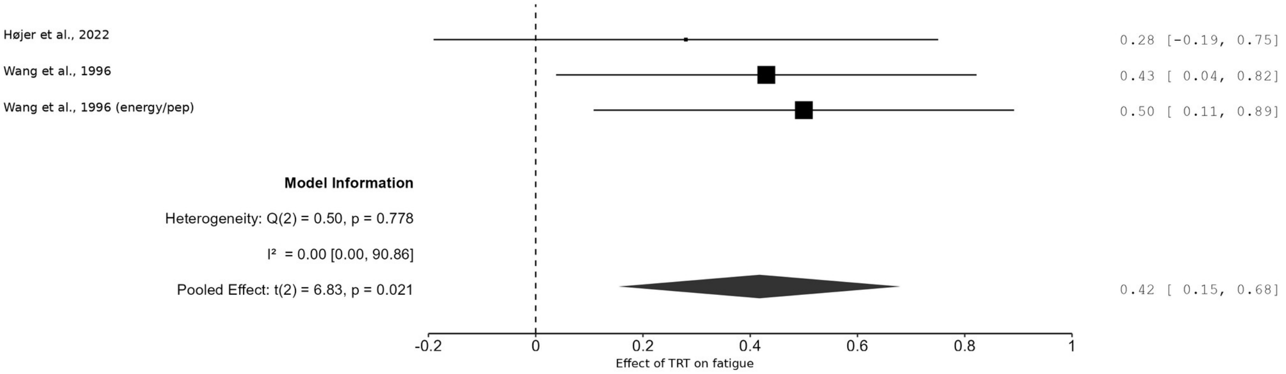


*Source: Authors, 2026.*

**Figure S4.** Forest plot of the effect of testosterone replacement therapy on erectile function in men with hypogonadism.

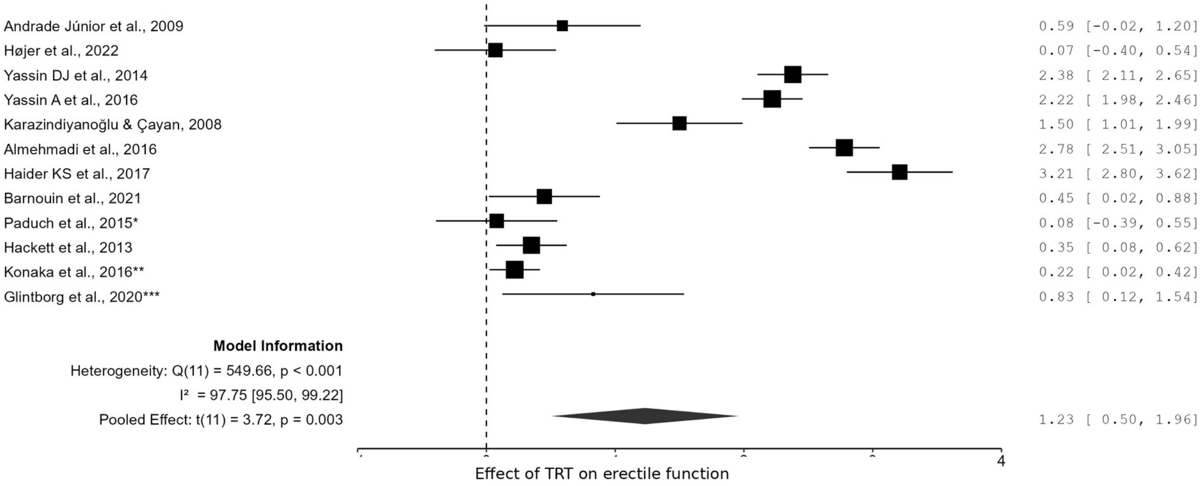


*Source: Authors, 2026.*

**Figure S5.** Forest plot of the effect of testosterone replacement therapy on fat mass in men with hypogonadism.

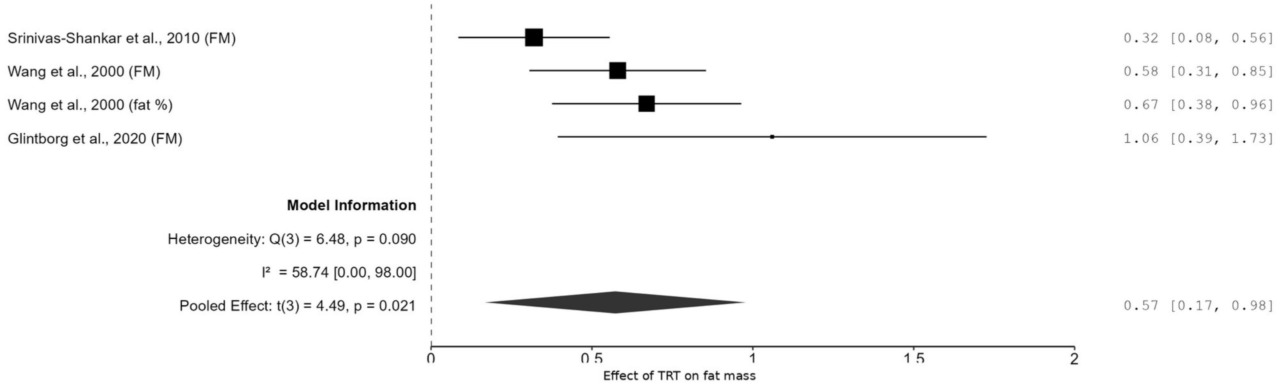


*Source: Authors, 2026.*

**Figure S6.** Forest plot of the effect of testosterone replacement therapy on lean mass in men with hypogonadism.

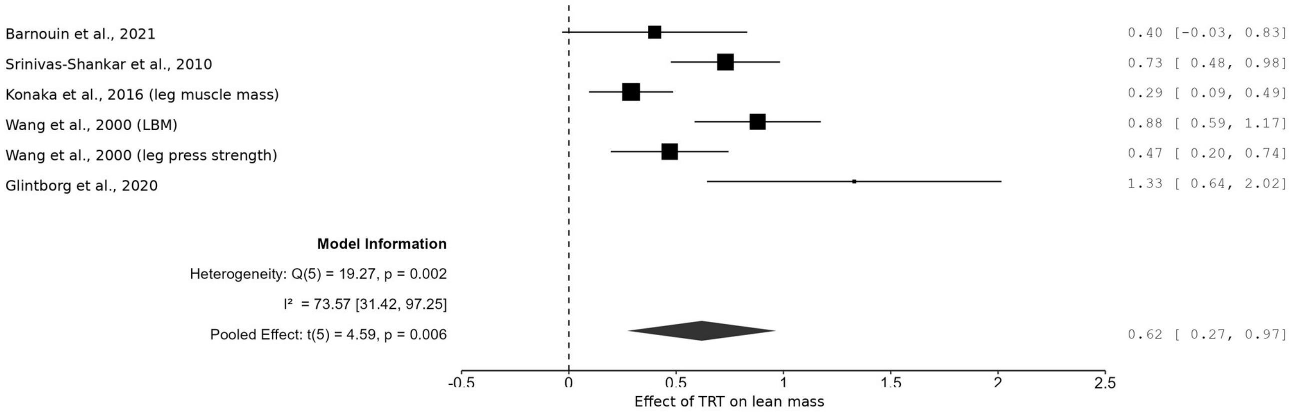


*Source: Authors, 2026.*

**Figure S7.** Forest plot of the effect of testosterone replacement therapy on quality of life in men with hypogonadism.

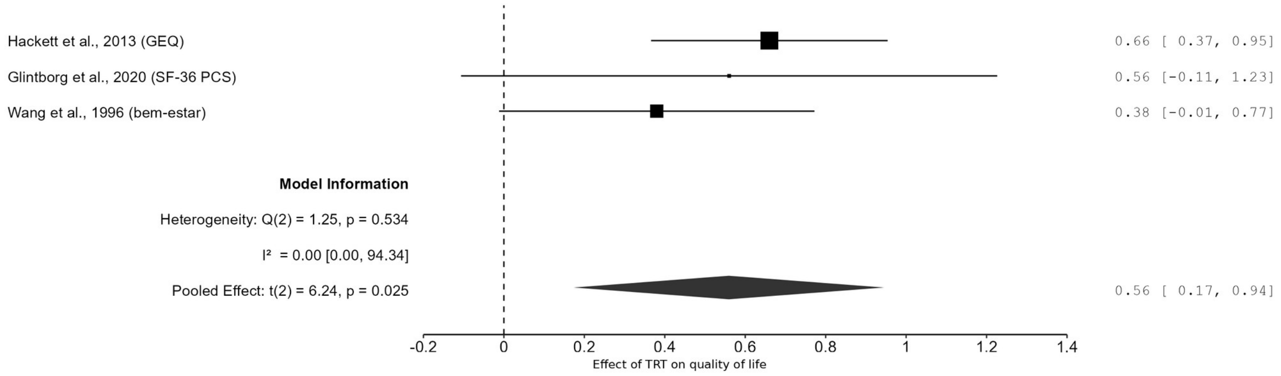


*Source: Authors, 2026.*

**Figure S8.** Forest plot of the effect of testosterone replacement therapy on hypogonadal symptoms assessed with the Aging Males' Symptoms (AMS) scale.

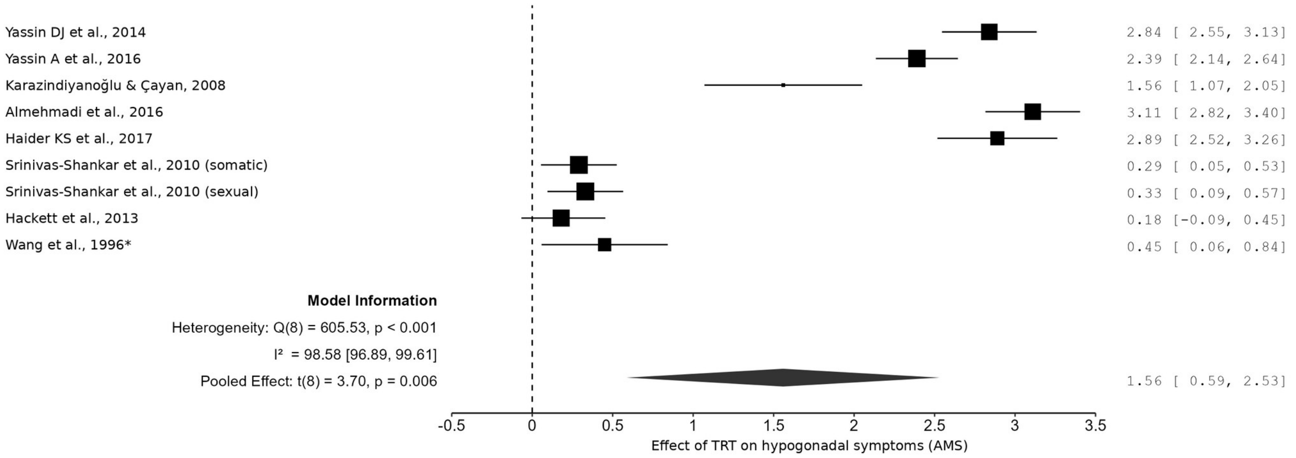


*Source: Authors, 2026.*

**Figure S9.** Forest plot of the effect of testosterone replacement therapy on lower urinary tract symptoms assessed with the International Prostate Symptom Score (IPSS).

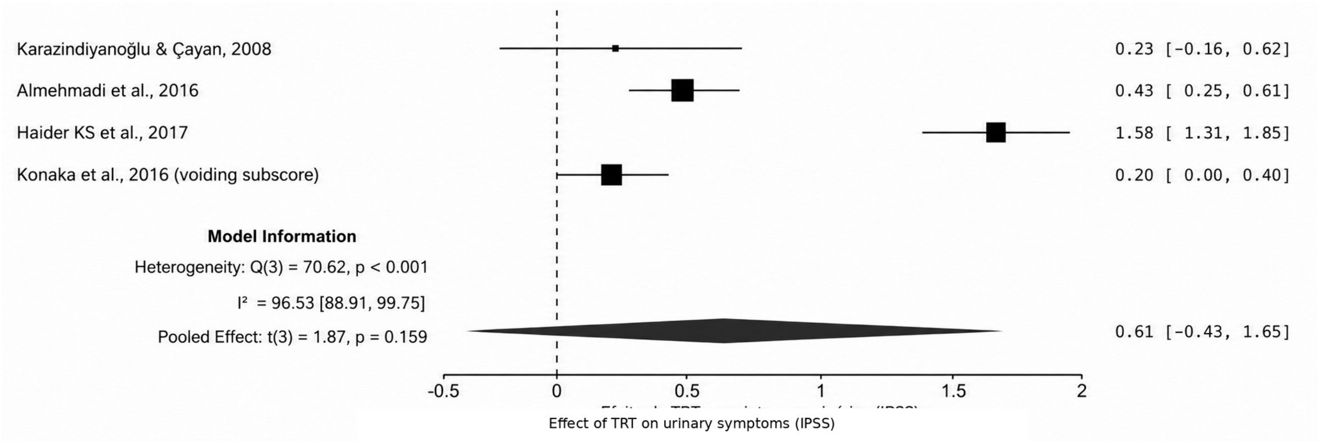


*Source: Authors, 2026.*
